# Prevalence of non-communicable disease risk factors and health-promoting lifestyle profiles, and Associations of nursing students in selected Schools of Nursing in the Western Province, Sri Lanka

**DOI:** 10.1101/2025.04.25.25326095

**Authors:** Dona Samantha Vajiramali Attygalle, Dewarahandi Kavishka Madushan De Silva, Sudath Shirley Pathmasiri Warnakulasuriya

## Abstract

**Background:** Non-communicable diseases (NCDs) are a leading cause of morbidity and mortality worldwide, and lifestyle factors are important etiological factors. It is essential to be aware of the prevalence of NCD risk factors and health-promoting behaviors among nursing students because they are a future healthcare workforce responsible for promoting healthy lifestyles. This study assessed the prevalence of risk factors for NCDs, health-promoting lifestyle behaviors, and their relationships among nursing students.

**Method:** A descriptive cross-sectional study was conducted in two randomly selected Schools of Nursing in the Western Province, Sri Lanka. Using a census sampling technique, 603 nursing students were enrolled. Data were collected through a self-administered questionnaire with three sections: socio-demographic information, NCD risk factors, and the Health-Promoting Lifestyle Profile II (HPLP II). Anthropometric measurements were taken to assess Body mass index(BMI). Data analysis was performed using SPSS statistical package (version 24), employing descriptive statistics, Pearson correlation, t-tests, ANOVA, and Chi-square rest.

**Results:** Among the participants, 78.9% were aged 23-25 years, and 93.4% were female. The prevalence of overweight and obesity was 11.4%, with males exhibiting a higher prevalence (21.62%) than females (10.89%) (p<0.05). Low physical activity was reported by 71.96% of students, with a higher prevalence among females (73.42%) than males (51.35%) (p<0.05). The overall HPLP II mean score (2.39±0.30) indicated a middle level of health-promoting behaviors, with low physical activity (1.87±0.49) and health responsibility (2.11±0.40) subscales scoring weakest. Significant correlations were found among all HPLP II subscales. Age, gender, and income were significantly associated with health-promoting behaviors.

**Conclusions:** Nursing students in the Western Province, Sri Lanka, have an alarmingly high rate of NCD risk factors, particularly low physical activity. Health-promoting behavior is moderate with some areas requiring improvement. Physical activity and health responsibility must be addressed through interventions to equip future nurses with healthy habits and enhance their role as health promoters.

**Key Message:** *What is already known on this topic:* Non-communicable diseases such as ischemic heart disease, diabetes, and hypertension are increasingly prevalent in Sri Lanka, driven by lifestyle changes including physical inactivity and unhealthy diets. Alarming trends among young adults, such as rising obesity rates and poor dietary habits, are particularly concerning.

*What this study adds:* This study provides updated, localized data on the prevalence of NCD key risk factors and health-promoting behaviors among nursing students in Sri Lanka who were underrepresented in previous studies. It highlights specific areas of concern, such as low physical activity and poor health responsibility, and identifies demographic factors associated with health behavior patterns.

*How might this study affect research, practice, or policy:* These findings can inform the development of targeted interventions in nursing education to promote healthier lifestyles among students. It also underscores the need to incorporate structured health promotion and physical activity programs within nursing curricula. On a policy level, it could support the implementation of wellness initiatives in healthcare training institutions to foster a healthier future workforce.

## Introduction

Non-communicable diseases (NCDs), including cardiovascular diseases, diabetes, cancer, and chronic respiratory diseases, are among the leading causes of morbidity and mortality worldwide [1]. It has been reported that approximately 80% of all deaths worldwide are attributed to NCDs, with low- and middle-income nations bearing a disproportionately high burden in 2024 [1]. Modifiable behavioral risk factors such as tobacco use, excessive alcohol use, poor diet, and physical inactivity are the leading causes of the increased prevalence of NCDs [2,3]. Further, metabolic risk factors, including obesity, hypertension, dyslipidemia, and hyperglycemia, also contribute to the increasing burden of these diseases [2,4,5]. Mitigating the long-term effects of NCDs on individuals and healthcare systems requires early detection and lifestyle modification of these risk factors.

Nursing students constitute a vital demographic within the healthcare system, as they are prospective frontline healthcare professionals [6]. The knowledge of these students, their perspectives, and individual health practices significantly impact their capacity to instruct and affect the patients concerning healthy living decisions [6]. However, previous studies have indicated that nursing students, like the general populace, may be susceptible to adopting unhealthy behaviors, such as inadequate dietary practices, sedentary lifestyles, and elevated stress levels[7–9]. According to Monterio [10], among the sampled nursing students, 65.1% consumed alcohol, 57.4% did not do physical activity, and 34.7% were overweight. Supporting the previous findings, Gamelen [11] also reported that among the nursing students, 28.6% of males and 30.4% of females were overweight, and more than half of them were obese. Surprisingly, one-third of female students (33.7%) were classified as prehypertension, and both males and females were physically inactive (71.4% and 81.2%)[7]. The high demands of nursing education, encompassing challenging academic courses, clinical rotations, and shift work, may lead to poor lifestyle behaviors among students [12,13]. Thus, identifying the prevalence of risk factors for NCDs and the health-promoting lifestyles of nursing students are crucial for creating focused interventions to enhance their health and, in turn, their capacity to promote healthy practices when providing patient care.

Health-promoting lifestyle behaviors encompass multiple dimensions, including physical activity, nutrition, stress management, interpersonal relationships, and self-care [14,15]. The Health-Promoting Lifestyle Profile (HPLP) is a commonly used framework for assessing individuals’ engagement in these habits and identifying developmental opportunities [14]. According to research, nursing students frequently engage in suboptimal health-promoting behaviors, with inadequate physical exercise and high levels of stress being the most prevalent concerns (Alzahrani et al., 2021). Nursing students in Iraq reported that their health-promoting lifestyle profile was significantly lower (score=123.2 ± 19.9) [8]. Physical activity scores were reported as extremely poor among these students (16.1 ± 4.9 out of 36). Given that nurses’ act as role models for their patients, nursing students must acquire and maintain healthy living habits for their own well-being and professional obligations.

According to previous studies, gender, academic year, socioeconomic status, and stress level all have a substantial impact on students’ health habits and risk factor profiles [16] [8][7]. However, very few studies have been undertaken on this topic concerning nursing students in Sri Lanka or similar settings in South Asia. Identifying the prevalence of these risk factors and their interrelationships might help policymakers, educators, and healthcare organizations to establish health promotion programs that are targeted to nursing students’ unique requirements. In this light, the current study aimed to assess the prevalence of NCD risk factors and examine the health-promoting lifestyle profiles and their associations among nursing students in selected Schools of Nursing.

## Materials and Methods

### Study Design and Setting

A descriptive cross-sectional study was conducted to assess the prevalence of NCD risk factors, health-promoting behaviors, and their associations among nursing students from March 2023 to September 2023. The current study was conducted in two randomly selected Schools in Nursing in Western Province, Sri Lanka. Out of 18 Schools of Nursing in Sri Lanka, four are located in the Western province. Western province is the most urbanized and populous region in the country. This area is likely to reflect diverse socio-economic backgrounds and lifestyle patterns, making it a relevant setting for studying NCD risk factors and health behaviors among future healthcare professionals. Random selection of two schools helped to reduce selection bias while maintaining the quality and depth of data collection. The Ministry of Health in Sri Lanka oversees a network of Schools of Nursing dedicated to educating and preparing nursing professionals across the country. These schools offer comprehensive nursing education programs, combining theoretical instruction with practical clinical training.

### Study Population

All nursing students in the selected two Schools of Nursing were the population for the current study.

#### Inclusion and Exclusion Criteria

All nursing students enrolled in the program were included in the study, regardless of their age, gender, or study level. Nursing students who were absent on the date of data collection were excluded from the study.

### Sample size and sampling technique

The current study employed a census sampling method, which enabled all nursing students in the two selected Schools of Nursing to be included in the study. Therefore, 603 nursing students were taken as the sample.

### Study instrument

The data were collected through a self-administered questionnaire, which consisted of three sections. Section 1 consisted of five items dedicated to collecting the socio-demographic data of the participants, including age, gender, monthly income, type of accommodations, and religion.

Section 02 assessed the NCD risk factor profiles of the participants through five items, which were developed by the researchers with the help of previous literature [17–29]. They were mainly focused on major risk factors for NCDs, including physical inactivity, alcohol use, cigarette smoking, fruit and vegetable consumption, and BMI. These factors were the most prevalent among nursing students according to previous studies [30–32].

Section 03 comprised a culturally adapted and validated Sinhala version of “Health–Promoting Lifestyle Profile “ (HPLP II), which is dedicated to measuring the health-promoting behaviors of the participants [33]. The original HPLP II questionnaire was developed by Walker [34] within a wellness framework and consists of 52 items designed to assess health-promoting behaviors. The HPLP II evaluates the frequency of engagement in health-promoting activities using a four-point Likert scale (1 = Never, 2 = Sometimes, 3 = Often, 4 = Routinely), with higher scores indicating more frequent health-promoting behaviors. The total score ranges from 52 to 208, with the highest possible score being 252. The questionnaire is categorized into six subscales: Health Responsibility (9 items), Physical Activity (8 items), Nutrition (9 items), Spiritual Growth (9 items), Interpersonal Relations (9 items), and Stress Management (8 items).

The Sinhala version of the HPLP-II demonstrated strong reliability and validity. Internal consistency was high, with a Cronbach’s alpha of 0.98, indicating that the instrument reliably measures health-promoting behaviors across its subscales. Additionally, the test-retest reliability was robust, with an Intraclass Correlation Coefficient (ICC) of 0.98 (95% CI = 0.97–0.99), confirming its stability over time. In terms of validity, the structural validity of the tool was confirmed through factor analysis using principal component analysis. The analysis revealed seven factors that explained 80.65% of the cumulative variance, supporting the tool’s ability to capture various dimensions of health-promoting behaviors.

### Data collection

After obtaining the ethical clearance from the Ethics Review Committee of KAATSU University, Battaramulla, data collection was commenced under the relevant permission from the two principals of the two selected Schools of Nursing. Two separate full days were allocated for two schools, and initially, the information sheets were provided, and the consent forms were obtained from the potential participants. Afterward, the printed questionnaire was distributed, and once they finished, height and weight were measured with the help of trainee data collectors. The participants’ height was measured using a calibrated stadiometer. The participants were asked to stand straight, with their heels together, and head in the Frankfort plane. The height was recorded in centimeters (cm). Participants’ weight was measured using a digital weighing scale. The participants were instructed to stand still on the scale, without shoes, and to wear minimal clothing. Weight was recorded in kilograms (kg).

All the information was gathered anonymously, and the collected data forms were kept under lock and key. The electronic data was password protected, and only investigators had access. All collected data will be stored for up to 5 years.

### Data analysis

The data were initially coded and entered into a Statistical Package for the Social Sciences (SPSS) version 24. Data were descriptively analyzed for frequencies, percentages, means, and standard deviations (SD). The association between gender and the prevalence of risk factors was generated through a chi-square test. The total score means and item means were separately generated for the HPLP II subscales. Health-promotive lifestyle was then categorized into three levels using item means. The lower third of the scale (1 - 2.33) was categorized as “week”, the middle third of the scale (2.34 - 3.33) was categorized as “Middle”, and the upper third of the scale (3.34 – 4.00) was categorized as “Good” health promotive behavior [35].

The Pearson correlation test generated the correlation between the subscales of HPLP II. The associations between HPLP II scores and socio-demographic characteristics were generated through independent sample t-tests and ANOVA tests. The significance threshold was set as p<0.05. Body mass index (BMI) was calculated by dividing a person’s weight by kilograms per square meter (kg/m2) and the BMI score was divided into four levels according to WHO guidelines as; underweight (< 18.5); normal, weight (18.5–24.9); overweight (25–29.9); and obese (= 30) [29].

### Findings

Out of the 603 nursing students from the two selected Schools of Nursing, 560 students (92.86%) responded.

### Socio-demographic characteristics of nursing students

The basic socio-demographic characteristics of the participants are presented in Table 1. Of the participants, the majority (n=442, 78.9%) were in the 23-25 age group. Gender distribution showed a significant female predominance, with 93.4% (n=523) of the participants being female. The vast majority of students identified as Buddhist (n=543, 97%). Regarding income, most students reported a monthly income between LKR 30,000 and 34,999 (n=474, 84.6%), with a smaller proportion earning more than LKR 40,000 (n=35, 6.3%). Accommodation patterns revealed that the largest group of students lived in hostels (n=286, 51.1%)

**Table 1.** Socio-demographic characteristics of the nursing students (n=560)

| Characteristics | Frequency (n) | Percentage (%) |
| --- | --- | --- |
| **Age (Years)** |  |  |
| 19-22 | 13 | 2.3 |
| 23-25 | 442 | 78.9 |
| 26-28 | 103 | 18.4 |
| 29-31 | 02 | 0.4 |
| **Gender** |  |  |
| Male | 37 | 6.6 |
| Female | 523 | 93.4 |
| **Religion** |  |  |
| Buddhist | 543 | 97 |
| Hindu | 08 | 1.4 |
| Catholic | 09 | 1.6 |
| **Income (LKR)** |  |  |
| 30,000 – 34,999 | 474 | 84.6 |
| 35,000 – 40,000 | 51 | 9.1 |
| >40,000 | 35 | 6.3 |
| **Accommodation** |  |  |
| Own House | 91 | 16.3 |
| Boarding location | 178 | 31.8 |
| Hostel | 286 | 51.1 |
| Other | 05 | 0.9 |

### Prevalence of NCD risk factors among nursing students

Sex-specific prevalence of the key NCD risk factors is summarized in Table 2. The prevalence of overweight and obesity was 11.4% (95% CI: 8.81–14.09) and males had a higher prevalence of overweight and obesity (21.62%, 95% CI: 8.36–34.89) compared to females (10.89%, 95% CI: 10.68–10.74) (p<0.05). The highest prevalence risk factor was observed for low physical activity 71.96% (95% CI: 68.24–75.68) of the participants. Low physical activity was significantly more common among females (73.42%, 95% CI: 69.64–77.21) than males (51.35%, 95% CI: 35.25– 67.46) (p<0.05). Active alcohol consumption was reported in 5.18% (95% CI: 3.34–7.01) of the total sample and was significantly higher among males (32.4%, 95% CI: 17.35–47.52) compared to females (3.25%, 95% CI: 1.73–4.77) (p<0.05).

**Table 2.** Prevalence of NCD risk factors among participants.

| Risk Factors | Male |  |  | Female |  |  | Total |  |  |
| --- | --- | --- | --- | --- | --- | --- | --- | --- | --- |
|  | No. | % | CI | No. | % | CI | No. | % | CI |
| <sup>1</sup>Overweight and obese * | 08/37 | 21.62 | (8.36-34.89) | 57/523 | 10.89 | (10.68-10.74) | 64/560 | 11.4 | (8.81-14.09) |
| <sup>2</sup>Low Physical activity * | 19/37 | 51.35 | (35.25-67.46) | 384/523 | 73.42 | (69.64-77.21) | 403/560 | 71.96 | (68.24-75.68) |
| Active smoking | 01/37 | 2.7 | (0.0 – 7.93) | 04/523 | 0.76 | (0.0 - 1.51) | 05/560 | 0.89 | (0.11-1.67) |
| Active alcohol consumption* | 12/37 | 32.4 | (17.35-47.52) | 17/523 | 3.25 | (1.73-4.77) | 29/560 | 5.18 | (3.34-7.01) |
| <sup>3</sup>Low fruit and vegetable consumption | 29/37 | 80.56 | (67.63-93.48) | 460/523 | 88.46 | (85.72-91.21) | 489/560 | 87.95 | (85.24-90.66) |

\*p<0.05 for male-female comparison CI=Confidence interval
<sup>1</sup> BMI level > 25kg/m<sup>2</sup>
<sup>2</sup> <150 minutes of moderate-intensity physical activity per week
<sup>3</sup> Consume < 2-4 servings of fruit each day and < 3-5 servings of vegetables each day

### Health-promoting lifestyle profile of nursing students

Health-promoting lifestyle profiles of the nursing students are presented in Table 3. The overall mean score of HPLP II was 126.52 (±16.37) and its item mean was 2.39 (±.39), which falls under the middle level of health promotion behavior. The Health Responsibility subscale had a mean score of 19.07 (±3.63), with an item mean of 2.11 (±0.40), indicating a weak level of engagement. Similarly, Physical Activity scored a mean of 15.03 (±3.94), with an item mean of 1.87 (SD = 0.49), also reflecting a weak level of activity. Other subscales, including nutrition, spiritual growth, interpersonal relationships, and stress management, fall under the middle level of health-promotive behaviors.

**Table 3.**
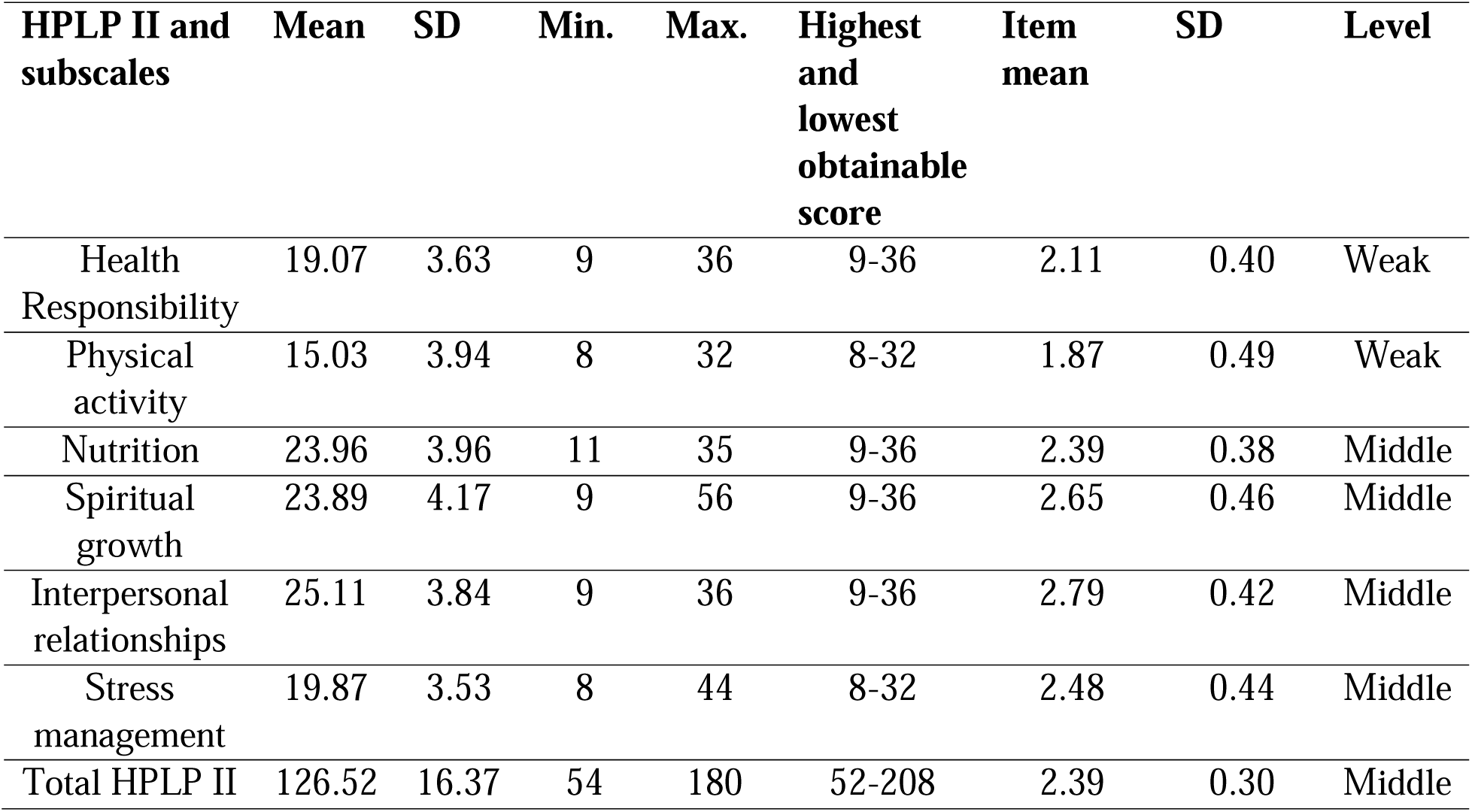
Health-promoting lifestyle profile of nursing students (n=560)

### Correlation between subscales

Table 4 presents the correlation coefficients between the subscales of the Health-Promoting Lifestyle Profile II (HPLP II). Significant positive correlations were observed across all subscales (*p*<0.05).

**Table 4.** Correlation between HPLP II subscales.

| **Subscales** | **Health Responsibility** | **Physical activity** | **Nutrition** | **Spiritual growth** | **Interpersonal relationships** | **Stress management** |
| --- | --- | --- | --- | --- | --- | --- |
| Health Responsibility<br>y<br>(r=) | 1 | 0.411** | 0.421** | 0.436** | 0.397** | 0.379** |
| Physical activity<br>(r=) | 0.411** | 1 | 0.304** | 0.314** | 0.231** | 0.369** |
| Nutrition<br>(r=) | 0.421** | 0.304** | 1 | 0.304** | 0.351** | 0.231** |
| Spiritual growth<br>(r=) | 0.436** | 0.314** | 0.304** | 1 | 0.623** | 0.557** |
| Interpersonal relationships<br>(r=) | 0.397** | 0.231** | 0.351** | 0.623** | 1 | 0.525** |
| Stress management<br>(r=) | 0.379** | 0.369** | 0.231** | 0.557** | 0.525** | 1 |

\*\* Correlation is significant at the 0.01 level (2-tailed)

### Associations between HPLP II scores and socio-demographic characteristics

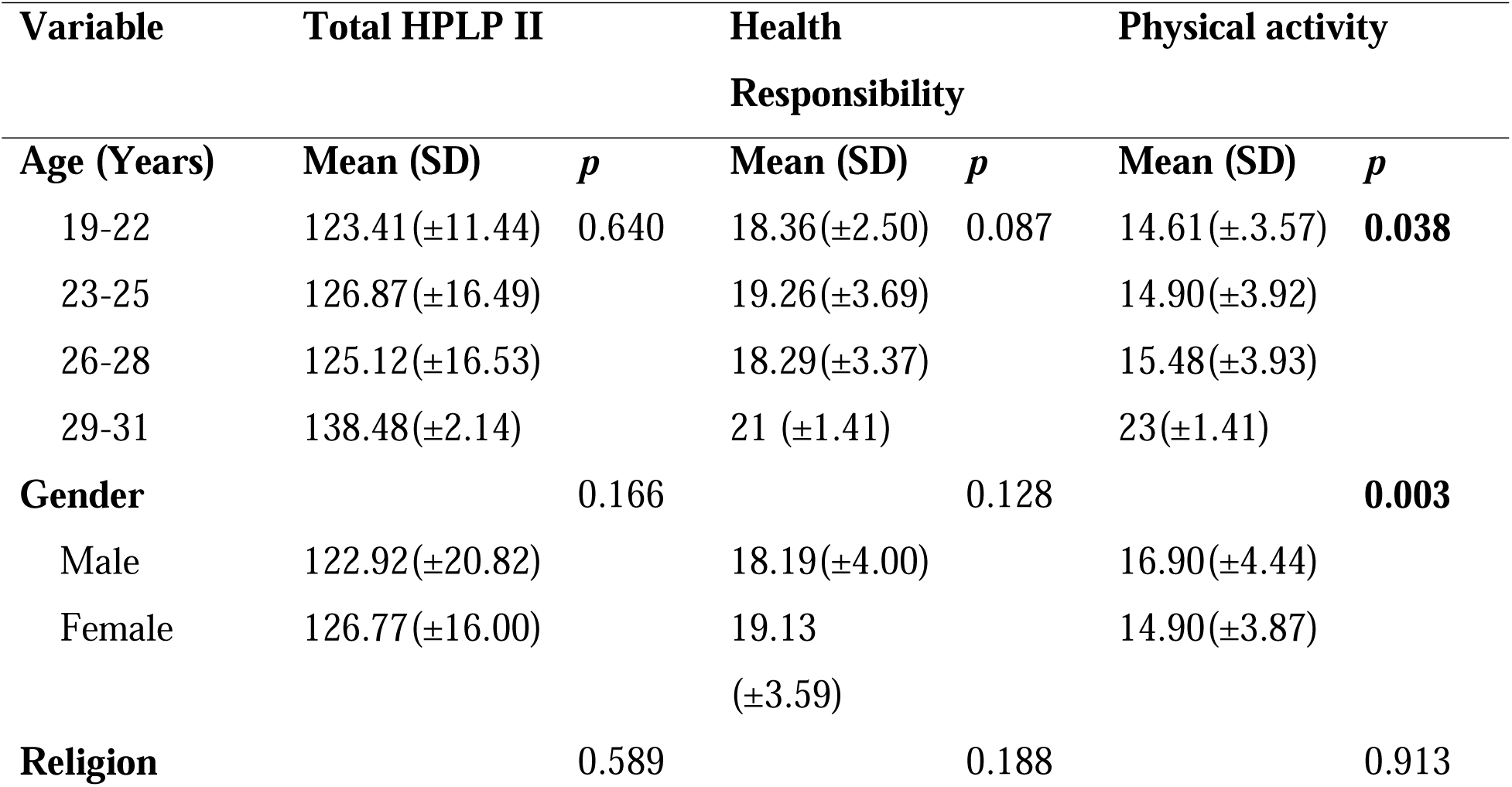
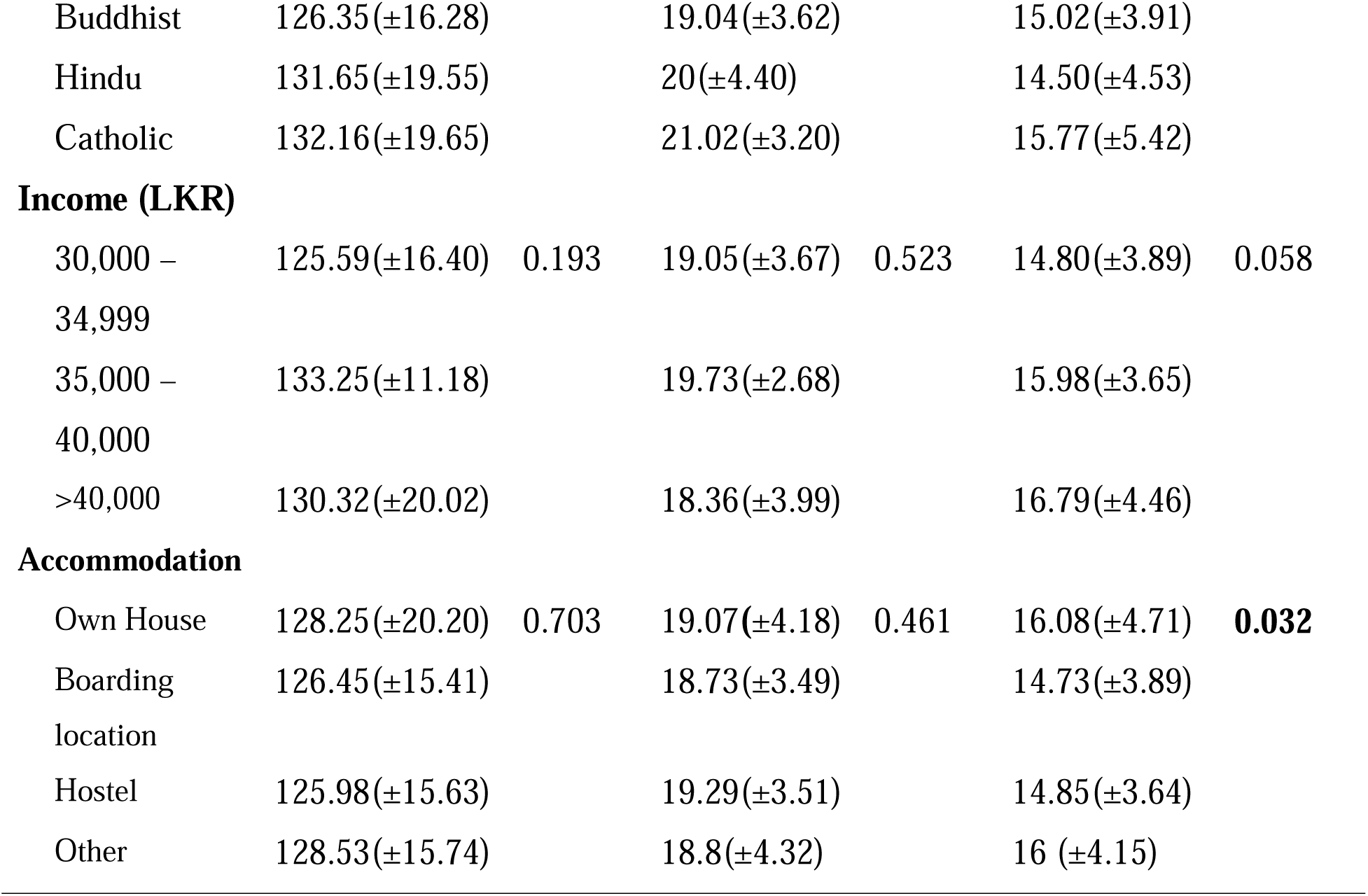

| Variable | Total HPLP II |  | Health Responsibility |  | Physical activity |  |
| --- | --- | --- | --- | --- | --- | --- |
| Age (Years) | Mean (SD) | *p* | Mean (SD) | *p* | Mean (SD) | *p* |
| 19-22 | 123.41(±11.44) | 0.640 | 18.36(±2.50) | 0.087 | 14.61(±3.57) | **0.038** |
| 23-25 | 126.87(±16.49) |  | 19.26(±3.69) |  | 14.90(±3.92) |  |
| 26-28 | 125.12(±16.53) |  | 18.29(±3.37) |  | 15.48(±3.93) |  |
| 29-31 | 138.48(±2.14) |  | 21 (±1.41) |  | 23(±1.41) |  |
| **Gender** |  | 0.166 |  | 0.128 |  | **0.003** |
| Male | 122.92(±20.82) |  | 18.19(±4.00) |  | 16.90(±4.44) |  |
| Female | 126.77(±16.00) |  | 19.13<br>(±3.59) |  | 14.90(±3.87) |  |
| **Religion** |  | 0.589 |  | 0.188 |  | 0.913 |
| Buddhist | 126.35(±16.28) |  | 19.04(±3.62) |  | 15.02(±3.91) |  |
| Hindu | 131.65(±19.55) |  | 20(±4.40) |  | 14.50(±4.53) |  |
| Catholic | 132.16(±19.65) |  | 21.02(±3.20) |  | 15.77(±5.42) |  |
| **Income (LKR)** |  |  |  |  |  |  |
| 30,000 – 34,999 | 125.59(±16.40) | 0.193 | 19.05(±3.67) | 0.523 | 14.80(±3.89) | 0.058 |
| 35,000 – 40,000 | 133.25(±11.18) |  | 19.73(±2.68) |  | 15.98(±3.65) |  |
| >40,000 | 130.32(±20.02) |  | 18.36(±3.99) |  | 16.79(±4.46) |  |
| **Accommodation** |  |  |  |  |  |  |
| Own House | 128.25(±20.20) | 0.703 | 19.07(±4.18) | 0.461 | 16.08(±4.71) | **0.032** |
| Boarding location | 126.45(±15.41) |  | 18.73(±3.49) |  | 14.73(±3.89) |  |
| Hostel | 125.98(±15.63) |  | 19.29(±3.51) |  | 14.85(±3.64) |  |
| Other | 128.53(±15.74) |  | 18.8(±4.32) |  | 16 (±4.15) |  |

Tables 5 and 6 represent the association between socio-demographic factors and HPLP II subscales. Age was significantly associated with physical activity, with individuals in the 19-22 years age group reporting lower physical activity scores (14.61 ± 3.57) compared to older groups, and males scoring higher in physical activity (16.90 ± 4.44) than females (14.90 ± 3.87) (p = 0.003). Income levels also significantly influenced nutrition scores, with those in the 35,000–40,000 LKR income group reporting the highest nutrition scores (25.08 ± 3.42) (p = 0.013). Additionally, gender was significantly associated with interpersonal relationship scores, as females reported higher scores (25.28 ± 3.73) than males (22.63 ± 4.56) (p = 0.001).

**Table 6.**
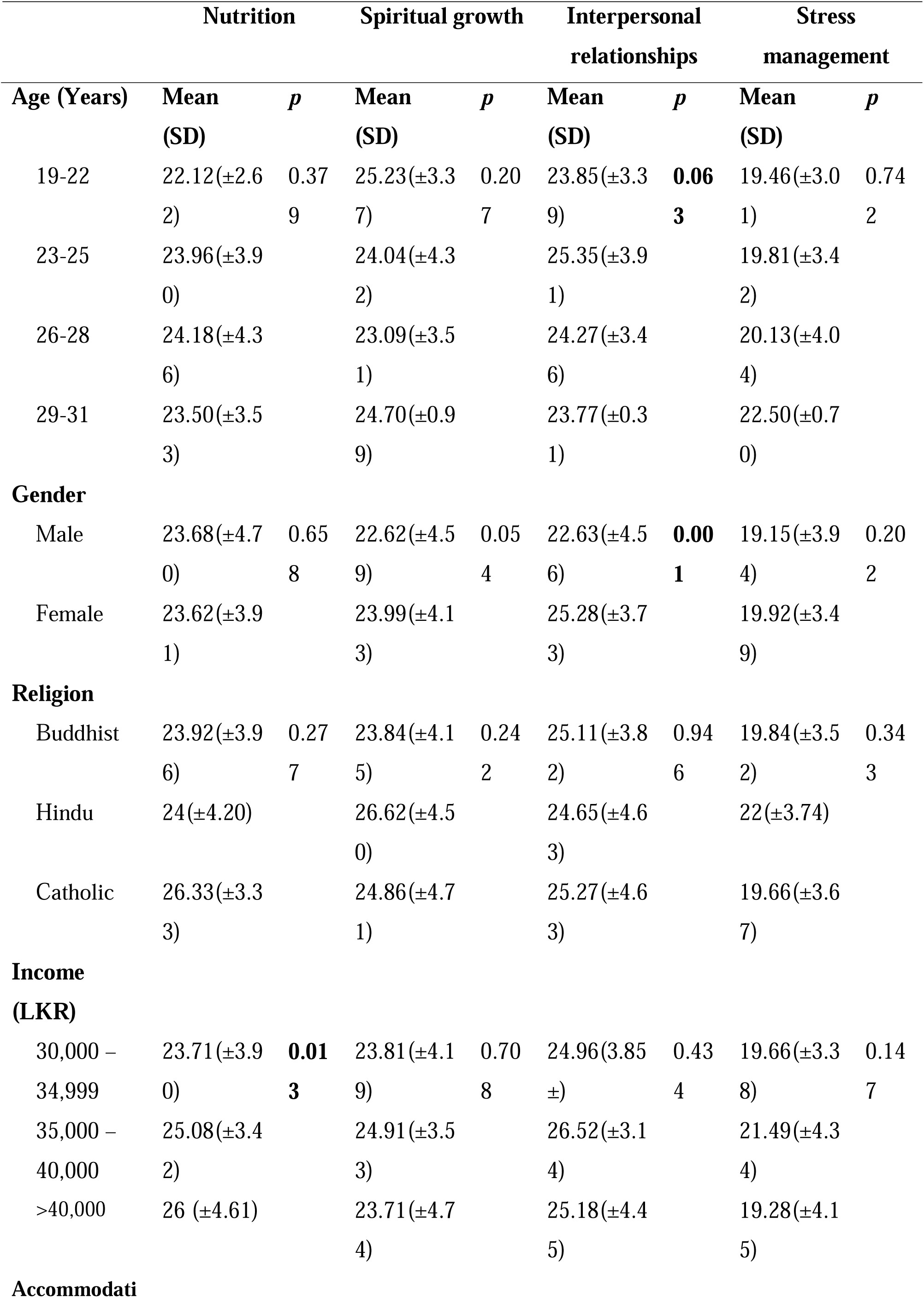
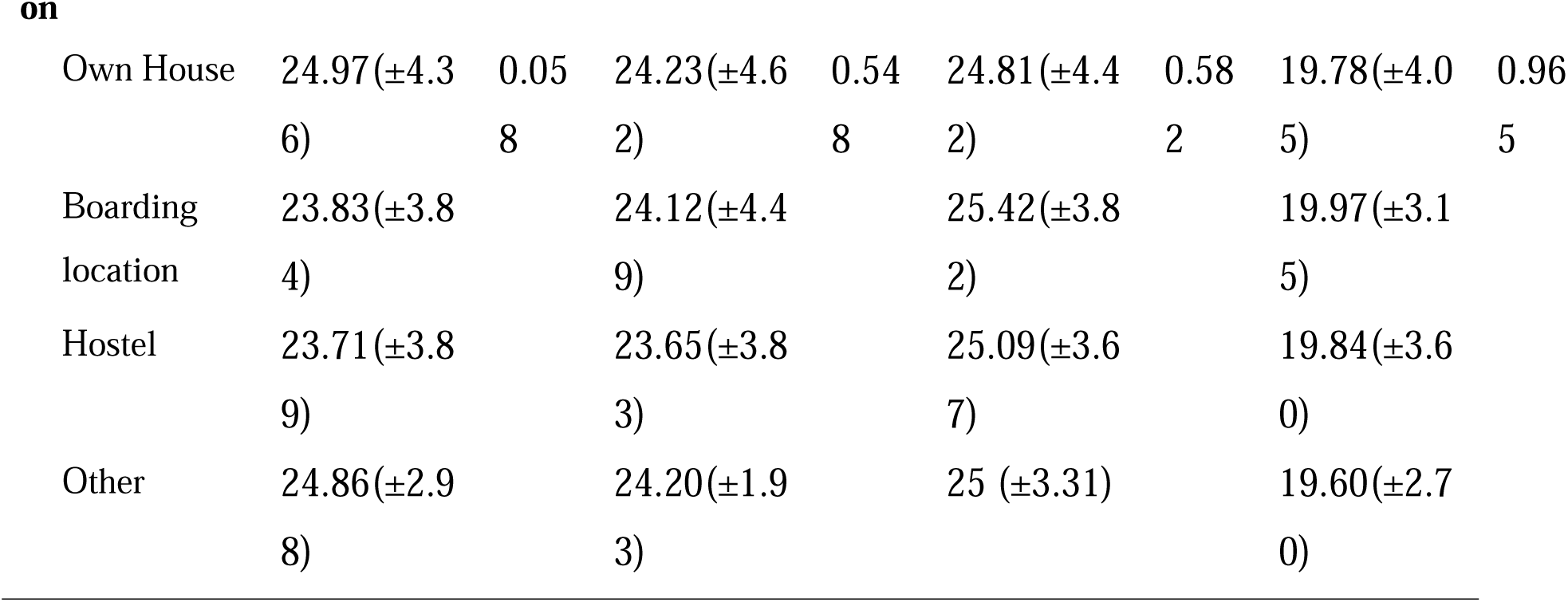
Association between socio-demographic factors and HPLP II subscales.

|  | Nutrition | Spiritual growth | Interpersonal relationships | Stress management |
| --- | --- | --- | --- | --- |

| Age (Years) | Mean<br>(SD) | *p* | Mean<br>(SD) | *p* | Mean<br>(SD) | *p* | Mean<br>(SD) | *p* |
| --- | --- | --- | --- | --- | --- | --- | --- | --- |
| 19-22 | 22.12(±2.6<br>2) | 0.37<br>9 | 25.23(±3.3<br>7) | 0.20<br>7 | 23.85(±3.3<br>9) | **0.06**<br>**3** | 19.46(±3.0<br>1) | 0.74<br>2 |
| 23-25 | 23.96(±3.9<br>0) |  | 24.04(±4.3<br>2) |  | 25.35(±3.9<br>1) |  | 19.81(±3.4<br>2) |  |
| 26-28 | 24.18(±4.3<br>6) |  | 23.09(±3.5<br>1) |  | 24.27(±3.4<br>6) |  | 20.13(±4.0<br>4) |  |
| 29-31 | 23.50(±3.5<br>3) |  | 24.70(±0.9<br>9) |  | 23.77(±0.3<br>1) |  | 22.50(±0.7<br>0) |  |
| **Gender** |  |  |  |  |  |  |  |  |
| Male | 23.68(±4.7<br>0) | 0.65<br>8 | 22.62(±4.5<br>9) | 0.05<br>4 | 22.63(±4.5<br>6) | **0.00**<br>**1** | 19.15(±3.9<br>4) | 0.20<br>2 |
| Female | 23.62(±3.9<br>1) |  | 23.99(±4.1<br>3) |  | 25.28(±3.7<br>3) |  | 19.92(±3.4<br>9) |  |
| **Religion** |  |  |  |  |  |  |  |  |
| Buddhist | 23.92(±3.9<br>6) | 0.27<br>7 | 23.84(±4.1<br>5) | 0.24<br>2 | 25.11(±3.8<br>2) | 0.94<br>6 | 19.84(±3.5<br>2) | 0.34<br>3 |
| Hindu | 24(±4.20) |  | 26.62(±4.5<br>0) |  | 24.65(±4.6<br>3) |  | 22(±3.74) |  |
| Catholic | 26.33(±3.3<br>3) |  | 24.86(±4.7<br>1) |  | 25.27(±4.6<br>3) |  | 19.66(±3.6<br>7) |  |
| **Income**<br>**(LKR)** |  |  |  |  |  |  |  |  |
| 30,000 –<br>34,999 | 23.71(±3.9<br>0) | **0.01**<br>**3** | 23.81(±4.1<br>9) | 0.70<br>8 | 24.96(3.85<br>±) | 0.43<br>4 | 19.66(±3.3<br>8) | 0.14<br>7 |
| 35,000 –<br>40,000 | 25.08(±3.4<br>2) |  | 24.91(±3.5<br>3) |  | 26.52(±3.1<br>4) |  | 21.49(±4.3<br>4) |  |
| >40,000 | 26 (±4.61) |  | 23.71(±4.7<br>4) |  | 25.18(±4.4<br>5) |  | 19.28(±4.1<br>5) |  |
| **Accommodati** |  |  |  |  |  |  |  |  |

|  |  |  |  |  |  |  |  |  |
| --- | --- | --- | --- | --- | --- | --- | --- | --- |
| Own House | 24.97(±4.3<br>6) | 0.05<br>8 | 24.23(±4.6<br>2) | 0.54<br>8 | 24.81(±4.4<br>2) | 0.58<br>2 | 19.78(±4.0<br>5) | 0.96<br>5 |
| Boarding<br>location | 23.83(±3.8<br>4) |  | 24.12(±4.4<br>9) |  | 25.42(±3.8<br>2) |  | 19.97(±3.1<br>5) |  |
| Hostel | 23.71(±3.8<br>9) |  | 23.65(±3.8<br>3) |  | 25.09(±3.6<br>7) |  | 19.84(±3.6<br>0) |  |
| Other | 24.86(±2.9<br>8) |  | 24.20(±1.9<br>3) |  | 25 (±3.31) |  | 19.60(±2.7<br>0) |  |

## Discussion

The findings reveal a significant age concentration in the 23-25-year age group, representing the majority of the participants, and the gender distribution in the study showed a striking female predominance, with more than nineteen percent of participants identifying as female. This is consistent with the trend observed in nursing education, where nursing has traditionally been a female-dominated profession globally, and this is reflected in the Sri Lankan context as well [36].

The current study identified several key NCD risk factors among nursing students, with significant gender differences. Concerning the overall prevalence, a minimal number of nursing students were overweight or obese in the current context. This amount is relatively low when compared to Sri Lankan community-based estimates that have seen increasing trends in body mass index (BMI) over the years [24, 25]. This may be possible because nursing programs emphasize health and wellness education, encouraging students to adopt healthier lifestyles. In contrast, broader community-based trends may reflect more sedentary lifestyles, poor dietary habits, and limited access to healthcare resources, leading to higher rates of overweight and obesity in the general population [25]. However, the prevalence of overweight and obesity was notably higher among male nursing students compared to female students. This finding supports the global trends where males are more likely to exhibit higher BMI levels [37]. In line with these findings, Bhagyalaxmi [26] reported that 30.8% of urban Indian males were overweight when compared to females (29.7%). But in contrast, Lopes [23] reported that among the Brazil university student’s prevalence of being overweight was higher in females (33.3%) than males (23.5%). Supporting these findings, another Brazilian study on students revealed the same pattern of higher prevalence of overweight among females [23]. This pattern is possible because men typically have more muscle mass and uneven fat distribution patterns [38], which impacts BMI measurements. Social and cultural influences also enter into the equation, with men possibly eating higher-calorie diets or not participating in weight-reduction programs to the same degree as women [22].

Despite a low prevalence of overweight, in the present study, obesity and the prevalence of inadequate physical activity are remarkably high, which warrants attention. However, the current findings broadly align with previous observations of sedentary lifestyles among South Asian populations [24, 25]. Interestingly, males are more adipose, and low physical activity levels are higher among female students (73.42%) than males (51.35%). In linear Lopes [23] identified that female students have a more sedentary lifestyle (60%) than males (53%). This finding is consistent with previous research that indicates women in South Asia have more restrictions in accessing physical exercise due to traditional roles, safety concerns, and a lack of recreational activities [21].

The fact that low levels of physical activity are predominantly found in females highlights a significant intervention area. Promotion of physical activity among both men and women is a critical aspect of the fight against NCDs since physical exercise is not only helpful in weight control but also has a significant role in reducing disease risks such as diabetes, hypertension, and cardiovascular disease [21]. These findings underscore the value of targeted interventions to enhance physical activity, particularly in female nursing students, and education regarding the value of active living for long-term health.

Low consumption of fruits and vegetables is also a concern, with the majority of students not meeting the recommended servings. This is a significant concern in the context of NCD prevention. Dietary factors play a significant role in the causality of NCDs, such as heart disease, diabetes, and certain cancers, and low intakes of fruits and vegetables have been an established risk factor for these conditions[18]. The finding is substantiated by systematic reviews in the field that have linked urbanization and lifestyle transition to poor diet [18,19]. In addition, Sri Lankan and other South Asian food cultures are more likely to revolve around calorie-dense and carbohydrate-dense foods [39], which may be less expensive and more accessible than fresh vegetables and fruits. Further, both sex groups are high in low fruit and vegetable intake, which is relatively higher in females because of variation in nutrition education, economic independence, and food cultural practices between Sri Lanka and the other South Asian context [18]. These findings suggest that a large proportion of nursing students are not meeting the nutritional standards necessary for optimal health, which increases their susceptibility to these chronic diseases over time.

Notably, the prevalence of active smoking and alcohol consumption is very low among nursing students. This prevalence is quite low when compared with global findings. According to Lopes [23], they reported that 10% of female and 40% of male students were smokers, while 63% of females and 100% of males were active alcohol drinkers in their context. These prevalence rates may reflect the social acceptability and availability of such substances for a predominantly young, educated population, which is consistent with other studies carried out with the Sri Lankan population [23,24]. Gender-wise, the prevalence of active smoking is 2.7% in males as opposed to 0.76% in females, and alcohol consumption is considerably higher in males compared to females. These differences reflect region-specific socio-cultural norms that discourage substance use among women while facilitating higher consumption among men [40].

To effectively address the prevalence of NCD risk factors among nursing students, it is crucial to recognize the role of health-promoting behaviors in mitigating these risks. By adopting and promoting these behaviors, nursing students can reduce their vulnerability to the identified risk factors, thereby enhancing their overall well-being and setting an example for future healthcare practices. Therefore, understanding and improving health-promotive behaviors is essential for mitigating the impact of NCD risk factors. The scores on the Health-Promoting Lifestyle Profile II (HPLP II) of the nursing students indicate various degrees of involvement in health-promoting practices across dimensions. In the Current study, the mean sum score of HPLP II indicated a mid-level practice of health-promoting activities among nursing students, which points towards an area for attention by future public health programs. This finding in this study suggests potential deficiencies in health education curricula and emphasizes the urgent need for implementing comprehensive health-promoting programs tailored for nursing students. Such interventions have proven effective in not only increasing students’ health awareness but also empowering them to adopt these behaviors into their everyday lives [41].

Considering the subscales, the “Physical activity” subscale yielded the lowest item mean score. This finding aligns with patterns observed in similar cohorts in other regions [42, 41]. Previous studies have shown that nursing students have low physical activity levels, leading to not just possible health drawbacks for the students themselves but also for the patients who receive care in the future [42,43].In line with the current findings, [42] reported that the physical activity subscale item mean score was 2.10 (±0.74) among nursing students in Korea. Similarly, an Iranian study also reported low physical activity behavior (2.69±0.80) among male nurses [35]. This pattern could be possible because sociological factors and academic pressure may be among the causes for this continued low level of physical activity among nursing students. However, the findings resonated with the need to intervene in structural obstacles to physical activity among nursing students.

The weak behavior pattern of physical activity level among nursing students is in line with the risk factor identified for NCDs because physical inactivity is a significant contributor to chronic diseases like heart disease, diabetes, and obesity. Physical inactivity contributes to poor cardiovascular health, weight gain, and other medical complications [17]. As low physical activity has a high correlation with NCDs, it is important that nursing students be encouraged to exercise often and be physically active to prevent such risks and improve well-being.

Similarly, the “Health Responsibility” subscale reported a low item mean, indicating a weak behavior pattern. The findings denoted that nursing students perceived less personal commitment to a health-oriented lifestyle. These findings align with global findings, which Dolu [44] also reported in their context that nursing students’ health responsibility is at a weak level mean score (2.18±0.52). This finding is especially alarming because nursing students are expected to act as role models for health-promoting behavior as future health professionals, as emphasized in the literature calling for nurses to practice the health behavior they preach [45]. When nursing students do not take care of their health, this raises doubts about their ability to promote the same practices for patients [46].

Conversely, the nutrition subscale indicates relatively higher adherence to healthy food habits, which are rated as middle-level. This concurs with the findings of research studies in other countries. According to Eshah [47] health-promoting behaviors reflect the same intermediate levels. However, the issue remains that although nutrition appears to be somewhat better, the overall way of life continues to require extreme modification.

The “Spiritual Growth,” “Interpersonal Relationships,” and “Stress Management” scales are scored at a middle level, reflecting sensitivity to the non-physical aspects of health. Given that nursing education is inherently stressful, avenues for improving interpersonal skills and effective stress management strategies directly impact students’ well-being and could be focal points for curricular revision to build resilience [48]. Interestingly, improved interpersonal relationships imply that the supportive system needed in these students may be enhanced through improved health outcomes [49].

The current findings further identified significant positive correlations between various subscales of the Health Promoting Lifestyle Profiles. Notably, health responsibility correlated strongly with physical activity (r = 0.411), nutrition (r = 0.421), spiritual growth (r = 0.436), interpersonal relationships (r = 0.397), and stress management (r = 0.379), indicating that the more health-responsible an individual is, the more likely he or she is to engage in a greater range of health-promoting behaviors. Physical activity also has high positive correlations with other subscales, suggesting that individuals who have a regular practice of physical activity might also have much appreciation for other health issues, such as nutrition and stress management. The strongest correlation was found between interpersonal relationships and spiritual growth (r = 0.623), indicating that individuals who focus on personal growth have improved social relations. These findings suggest that health-promoting behaviors are interrelated and that improvement in one dimension of health behavior can lead to improvement in other dimensions, contributing to overall well-being.

The findings of the current study further demonstrate significant associations between socio-demographic factors and health-protective behaviors among Sri Lankan nursing students, notably physical activity, interpersonal relationships, and nutrition. Notably, age was significantly related to physical activity, with the students in the age group of 29-31 years exhibiting higher levels of physical activity (p = 0.038). This suggests that older students may feel more accountable for their own health, probably due to they were more mature and had more life experience [50]. Younger students, particularly those in the age group 19-22 years, saw themselves as having low physical activity levels, which may reflect a lack of health behavior priorities in the early days of their studies. Gender differences were also significant, with female subjects scoring higher on the interpersonal relationship subscale value than male subjects. This reflects social norms whereby women are taught to attend to relational and emotional well-being and so engage in more healthy social acts [51]. In addition, income was also significantly associated with nutrition, where students from the 35,000 – 40,000 LKR income group had the highest scores on this subscale. Students from higher-income groups would likely have greater access to healthy foods, and this may result in healthier eating habits. This is in contrast to lower-income students, who would likely have money issues where they are limited from purchasing healthy foods, which would impact their nutritional habits [52]. These results highlight the complex interdependence of socio-demographic determinants affecting health behavior and suggest that age, gender, and income have significant implications in deciding lifestyle among nursing students. Strategies based on these could maximize health status among the population, for instance, increasing exercise among younger students, support systems among male students, nutrition guidance, and accessibility among low-income segments.

### Limitations

The current study has several limitations. A selected number of key NCD risk factors, which were highly reported in previous literature, were examined, which may have failed to capture the entire range of effects on health outcomes. The generalizability is limited since data were collected from two selected Nursing Schools. Data contamination cannot be precluded, which may impact the validity of the results. The use of a descriptive research design also limits the ability to establish causal relations between variables.

## Conclusion

In conclusion, the current study sheds light on several impressive findings related to NCD risk factors and health-promoting behaviors among nursing students in Sri Lanka. Prevalence of key NCD risk factors such as low physical activity, low fruit and vegetable consumption, and overweight/obesity was reported, and gender disparity differences were observed. Specifically, a higher prevalence of overweight/obesity was reported in males, while female subjects were less physically active and had a greater prevalence of low fruit and vegetable consumption. Additionally, current alcohol consumption was significantly prevalent among males compared to females. The overall health-promoting lifestyle profile of the students was in the middle level, with poor engagement in physical activity and health responsibility, while other areas such as nutrition, spiritual growth, interpersonal relationships, and stress management were in the middle level of health-promoting behaviors. Correlation analysis showed that there were significant positive correlations between all the subscales, indicating a holistic approach toward health promotion. Socio-demographic variables, including income, gender, and age, significantly affected health-promoting habits. Younger students indicated lower levels of physical activity, whereas higher-income groups of income had better nutrition scores. In addition, females reported higher scores in interpersonal relationships due to their increased participation in social well-being. These findings suggest the necessity of specific NCD risk factors and healthy lifestyle interventions among nursing students, namely physical activity and nutrition. Given the prime role of nursing students in health care provision, their health behavior improvement will not only contribute to their health but also have a beneficial influence on their subsequent health education and practice.

## Data Availability

All data produced in the present work are contained in the manuscript

## Patient and public involvement

Patients and/or the public were not involved in the design, conduct, reporting, or dissemination plans of this research.

## Patient consent for publication

Not required.

## Ethical approval and consent to participate

The current study was ethically cleared by the Ethics review committees established at Kaatsu Highly Advanced Medical Technology Training Centre (KIU/ERC/23/030). The study was conducted following the Declaration of Helsinki. Written informed consent was obtained from each participant before data collection.

## Availability of data and materials

All data relevant to the study are included in the article.

## Authors Contribution

SSPW and DSVA were responsible for conceptualizing the study and developing the methodology. SSPW supervised the project. All authors actively participated in the implementation of the study and the collection of data. DKMDS was responsible for conducting the formal data analysis and interpreting the results. The initial draft of the manuscript was collaboratively written by all authors. SSPW has revised the manuscript, organized its structure, and enhanced its content. Finally, all authors carefully reviewed and approved the final version of the manuscript, ensuring its accuracy and overall quality.

## Acknowledgments

The authors would like to express gratitude to all the Nursing students who participated in the study.

